# Cognitive features of amnestic Mild Cognitive Impairment using specific Cambridge Neuropsychological Test Automated Battery test scores

**DOI:** 10.1101/2022.06.09.22276176

**Authors:** Vinh-Long Tran-Chi, Arisara Amrapala, Gallayaporn Nantachai, Solaphat Hemrungrojn, Chavit Tunvirachaisakul, Michael Maes

## Abstract

**Background:** In older adults with amnestic Mild Cognitive Impairment (aMCI), the Cambridge Neuropsychological Test Automated Battery (CANTAB) probes indicated cognitive impairments most frequently in memory.

**Objectives:** This study aimed to investigate a) the cognitive features of aMCI using memory CANTAB tests and b) whether the clinical diagnosis of aMCI can be externally validated by these CANTAB measurements.

**Methods:** We tested CANTAB tests that are specific to aMCI on 65 healthy controls and 66 people with aMCI who were diagnosed using Petersen’s criteria. These tests were spatial working memory (SWM), visual pattern recognition memory (PRM), delayed matching to sample (DSM), spatial span (SSP), and rapid visual information processing (RVP).

**Results:** The key aMCI features are impairments in PRM and DSM, whilst deficits in SSP and RVP are other, albeit somewhat less important features of aMCI. Nevertheless, neural network analyses including 10 CANTAB domains specific for MCI showed that only 70.8 percent of all subjects were properly identified with a sensitivity of 77.3%, specificity of 65.4% and an area under the ROC curve of 0.760. K-means cluster analysis using the same specific CANTAB test scores discovered 2 clusters with an adequate silhouette measure of cohesion and separation including a cluster with 36 subjects showing impairments in most neurocognitive tests.

**Conclusion:** Deficits in spatial working, pattern recognition and visuospatial working memory as well as rapid visual information processing are key features of aMCI. Nevertheless, the clinical diagnosis of aMCI according to Petersen’s criteria is overinclusive because too many healthy controls are allocated to this group.

## Introduction

Mild cognitive impairment (MCI) affects a large part (10–15%) of older adults over the age of 65 (Anderson, 2019). Mild impairments in episodic memory, executive functions, visuospatial skills, processing speed, cognitive flexibility, and problem-solving ability are key features of aMCI, whilst basic activities of daily living (ADL) remain unaffected (Dwolatzky et al., 2004; Gualtieri & Johnson, 2005; Hemrungrojn et al., 2021; Petersen et al., 2010). aMCI may be regarded as a cognitive stage between healthy aging and dementia (Anderson, 2019), though the incidence of aMCI conversion to Alzheimer’s disease (AD) is roughly 16.5% each year and some aMCI patients (8%) recover from this condition (Petersen et al., 2010).

Sensitive and reliable neurocognitive tests are pivotal for the diagnosis of aMCI and AD, including the Montreal Cognitive Assessment (MoCA) and Mini-Mental State Examination (MMSE). The latter was developed in 1975 to evaluate dementia and neurocognitive deficits (Folstein et al., 1975), and has been used in the Thai population (Thai Cognitive Test Development Committee, 2002). Nasreddine et al. (2005) developed the MoCA as a rigorous and reliable MCI screening tool (Freitas et al., 2011). Regarding the ability to differentiate aMCI patients from those with healthy aging and dementia, the MoCA appears to have more accurate diagnostic properties than the MMSE. More precisely, the MoCA has a sensitivity and specificity of 90% and 100%, respectively, whilst the MMSE’s measures are 18% and 78%, respectively (Nasreddine et al., 2005). The MoCA has been translated and validated into Thai with high reliability and construct validity (Hemrungrojn et al., 2021). While normal controls and aMCI individuals may be adequately discriminated from AD patients using the MoCA total score, the MoCA subdomains, and other neuropsychological test results including the Consortium to Establish a Registry for Alzheimer’s Disease (Morris et al., 1988), the discrimination of aMCI from controls is more dubious (Tunvirachaisakul et al., 2018). Moreover, there is also some debate as to whether aMCI is a homogenous group of individuals or whether some of the aMCI subjects in fact belong to the normal control samples (Maes & Tangwongchai, 2021).

The Cambridge Neuropsychological Test Automated Battery (CANTAB) is a computerized program comprising different reliable neuropsychological tests, which is used to detect cognitive impairments in older adults with aMCI (Cotter et al., 2018; Roque et al., 2011). In older adults with aMCI, CANTAB probes have revealed cognitive impairment across a variety of areas, with memory functions being the most frequently impaired (Égerházi et al., 2007; Fowler et al., 2002; Johns et al., 2012; Robbins et al., 1998).

Working memory deficits are often found in aging individuals, especially those with MCI (Huntley & Howard, 2010; Saunders & Summers, 2010). Recent findings suggest that early impairments in visual episodic memory, executive function, semantic language/memory, attention, and working memory are strong predictors of progression from MCI to AD (Brandt et al., 2009; Klekociuk et al., 2014; Saunders & Summers, 2010). Kochan et al. (2011) reported that functional neuroimaging during a graded working memory task may help in the early detection of cognitive decline. Nevertheless, there is no data on whether CANTAB probes which assess memory could be used to externally validate the clinical diagnosis of aMCI employing supervised as well as unsupervised learning techniques.

Hence, the goals of our study were to investigate a) the cognitive features of aMCI using memory CANTAB tests; and b) whether the clinical diagnosis of aMCI may be externally validated by those CANTAB measurements (CANTAB, 1988), including spatial working (SWM, which probes “retention and manipulation of visuospatial information”) and visual pattern recognition memory (PRM), delayed matching to sample (DMS, which probes “short-term visual recognition memory”), and spatial span (SSP, which probes “visuospatial working memory capacity”) and rapid visual information processing (RVP). Since the clinical diagnosis aMCI is made using memory impairments, the a priori hypothesis of this study is that aMCI is accompanied by impairments in SWM, PRM, DMS, and SSP. Toward this end, we examine the differences in these CANTAB tests between aMCI individuals and controls using classical statistical tests as well as supervised learning techniques (binary regression analysis and neural networks). Furthermore, we examine whether unsupervised learning techniques with the CANTAB memory tests as input variables may retrieve the sample of clinically delineated subjects with aMCI according to Petersen’s criteria. If positive, the clinical diagnostic criteria of aMCI would be externally validated.

### Methods Participants

We conducted a cross-section study where aMCI participants were compared to healthy controls. Participants consisted of both sexes and had an age range of 55 to 84 years. Healthy volunteers were recruited from Pathumwan district, Bangkok, whilst aMCI patients were recruited from the Outpatient Department of King Chulalongkorn Memorial Hospital’s Dementia Clinic in Bangkok, Thailand. In older adults, the diagnosis of aMCI was made using the clinical Petersen’s criteria, namely: the presence of subjective and objective memory impairments and absence of dementia and changes in activities of daily living (ADL). In addition, aMCI individuals had a modified Clinical Dementia Rating (mCDR) score of 0.5 and controls a score of 0. Our normal controls included healthy older adult visitors of the Health Check-up Clinic, members of neighborhood senior clubs, healthy elderly caregivers of aMCI patients of the Dementia Clinic, and senior Red Cross volunteers.

Controls and aMCI individuals were not included in the study if they suffered from Alzheimer’s disease, or frontotemporal or vascular dementia or other types of dementia, major psychiatric disorders including substance use disorders, schizophrenia, major depressive disorders and bipolar disorder, abnormal kidney, liver and VDRL tests and vitamin B12 and abnormal thyroid concentrations, and neurodegenerative or neuroinflammatory disorders such as multiple sclerosis, stroke, Parkinson’s disease, trauma capitis, and meningitis. Finally, all subjects were assigned to one of two study samples, namely 65 healthy controls and 66 subjects with aMCI.

Prior to participation, all volunteers and aMCI subjects provided written informed consent. Our study adhered to both Thai and international ethical and privacy standards and is in accordance with the International Guideline for the Protection of Human Subjects, as required by the Declaration of Helsinki, the Belmont Report, the International Conference of Harmonization in Good Clinical Practice, and the CIOMS Guidelines. This study was approved by the Institutional Review Board (IRB) of the Faculty of Medicine, Chulalongkorn University, Bangkok, Thailand (No. 359/56).

### Clinical measurements

To determine the severity of aMCI, we used the Thai Mini-Mental State Examination (TMSE) and Thai Montreal Cognitive Assessment (MoCA-Thai). The TMSE is a Thai population-validated version of the MMSE, which consists of six fundamental subtests measuring: (1) orientation, (2) registration, (3) attention, (4) word recall, (5) language, and (6) computation. The total score ranges from 0 to 30. The TMSE is a test of global cognitive function that can be used to check for cognitive impairments. On the other hand, the MoCA may be used to distinguish between people experiencing typical age-related cognitive decline and those experiencing aMCI (Hemrungrojn et al., 2021). This test measures various cognitive domains, namely: (1) visuospatial, (2) executive, (3) language, (4) attention, (5) memory, and (6) orientation. The MoCA total score is calculated out of 30. Unlike the TMSE, the MoCA places greater importance on deficits of the frontal and executive function, as well as attention deficits. Additionally, it can also be used to discover neurocognitive dysfunctions within populations that have TMSE scores that fall in the normal range (Jirayucharoensak et al., 2019).

We used the CANTAB to measure neurocognitive performance tests deemed to be critical assessments for aMCI by CANTAB, namely Spatial Working Memory (SWM), Pattern Recognition Memory (PRM), Pattern Delayed Matching to Sample (DMS), Motor Planning Task (MOT), Spatial Span Length (SSP), and Rapid Visual Information Processing (RVP) (CANTAB, 1988). Retention and manipulation of visuospatial information in working memory is assessed using SWM. We used the SWM_BEr (SWM Between Error) and SWM_Str (SWM Strategy). PRM probes pattern recognition memory and we assessed PRM_Cor (PRM Correct), the subject’s total number of correct responses. The DMS assesses the subject’s ability to conduct simultaneous short-term visual recognition memory and visual matching. After a brief pause, a complicated visual pattern is presented to the subject and the subject must identify the matching pattern amongst the choice of four patterns. DMS_Cor (Total Correct) is the sum of trials where the individual has correctly responded to the initial stimulus, whilst DMS_MdCorL (Median Correct Latency). Another measure is the DMS_PEr (Percent Correct), which is the proportion of trials where the correct stimulus has been selected. MOT is used to assess gross motor function, namely motor speed, and movement precision. In this study, we employed the MOT_MdL (Median Latency), the median time required to respond during ten assessed trials. RVP assesses the capacity for prolonged visual attention. We assessed RVP_A′ and RVP_MdL (Median Latency), a measure that quantifies the subject’s effectiveness in detecting target sequences and quantifies the median response time, respectively. The subject’s capability to judge his or her own attention span and working memory was assessed using SSP. Here, the SSP_SpanL (Span Length), the longest sequence successfully recalled, was used.

### Statistics

We used IBM SPSS version 28 for Windows to conduct the statistical analysis. Two-tailed tests were applied and a statistical significance p-value of 0.05 was used. We employed analysis of variance to check differences in continuous variables across study groups and analysis of contingency tables (χ2-test) to check relationships between categorical variables. Multivariate general linear model (GLM) analysis was used to examine the relationships between diagnostic classifications and clinical and cognitive data after covarying for gender, age, and education. Assessment of univariate relationships between the classes and cognitive and clinical data was done by utilizing between-subject effects tests. Subsequently, the estimated marginal means (SE) were computed from the GLM model after adjusting for the gender, age, and education variables. To examine pair-wise differences in group means, we employed the protected least significant difference. Multiple comparisons were corrected for using the false discovery rate p-correction. We performed multiple regression analysis to determine which CANTAB test scores best predicted the MoCA and TMSE scores using a stepwise algorithm. For this analysis, we always confirmed multivariate normality, homoscedasticity, and the absence of multicollinearity. Additionally, 1000 bootstrap samples were used in the regression analysis, with the latter results being displayed if the results were not concordant. Clusters of individuals, based on the CANTAB test results, were constructed using K-means clustering and the two-step cluster analysis.

Partial least squares (PLS)-SEM analysis was conducted to examine the effects of the 10 CANTAB scores (input as single indicators) on a latent vector that was extracted from the instruments to make the clinical diagnosis of aMCI, namely Petersen’s criteria, MoCA and TMSE scores. Sex, age, and education could predict the latent vector and the CANTAB test scores. Complete PLS analysis was performed when the quality data of the model complied with the following criteria: (1) standardized root mean residual (SRMR) values < 0.08; (2) latent vectors display good convergence and construct validity as measured by an average variance extracted (AVE) > 0.5 and composite reliability > 0.7, factor loadings > 0.666 at p< 0.001; (3) the latent vector is not misspecified as a reflective model (as indicated by confirmatory tetrad analysis). PLS-SEM analysis was used to derive path coefficients (exact p values) as well as specific indirect and total effects, using 5000 bootstrap samples. The prediction power was evaluated using blindfolding and 10-fold cross-validation with PLSpredict.

## Results

### Clinical and demographic information

From the demographics and clinical features of the two study groups (**Table 1**), no significant differences were found in sex distribution or age between the groups. Additionally, we found that aMCI subjects had lower years of education than those with aMCI. The TMSE and MoCA scores were much lower in those with aMCI than healthy controls.

**Table 1.** Socio-demographic data of the participants in this study, divided into two groups: healthy controls (HC), and subjects with amnestic mild cognitive impairment (aMCI)

| Variables | HC (n=65) | aMCI (n=66) | F/ $\chi^2$ | df | p |
| --- | --- | --- | --- | --- | --- |
| Age – years (±SD) | 70.7 (±5.6) | 71.9 (±7.6) | 1.181 | 1/129 | 0.279<sup>a</sup> |
| Sex (F/M) | 57/8 | 63/3 | 2.565 | 1 | 0.109<sup>b</sup> |
| Education – years (±SD) | 13.6 (±4.6) | 10.0 (±5.3) | 17.152 | 1/129 | <0.001<sup>a</sup> |
| TMSE (±SD) | 28.9 (±1.1) | 27.0 (±1.8) | 54.039 | 1/129 | <0.001<sup>a</sup> |
| MoCA (±SD) | 26.6 (±1.6) | 20.1 (±2.8) | 264.06 | 1/129 | <0.001<sup>a</sup> |

*Note:* SD – Standard Deviation, <sup>a</sup> One-way ANOVA, <sup>b</sup> $\chi^2$ – test; F/ $\chi^2$ , Results of analyses of variance (F) or analyses of contingency analyses ( $\chi^2$ ); df, degree of freedom; p, p value.
*Abbreviations:* TMSE, Thai Mental State Examination; MoCA, Montreal Cognitive Assessment.

Multivariate GLM analysis that examined the connections between the outcomes of the CANTAB tests and the diagnosis, after controlling for age, education, and gender, is displayed in **Table 2**. A strong link between the CANTAB test results and the diagnosis was identified and significant influences from age and education were observed. The between-subject effects analyses revealed significant associations between diagnosis and all CANTAB test scores, except the RVP_MdL. These results were unaffected by adjusting for false discovery rates. Nevertheless, the effect sizes were <0.10 for all CANTAB test results. Each participant group’s computed model-generated marginal mean values for the CANTAB scores are shown in **Table 3**.

**Table 2.**
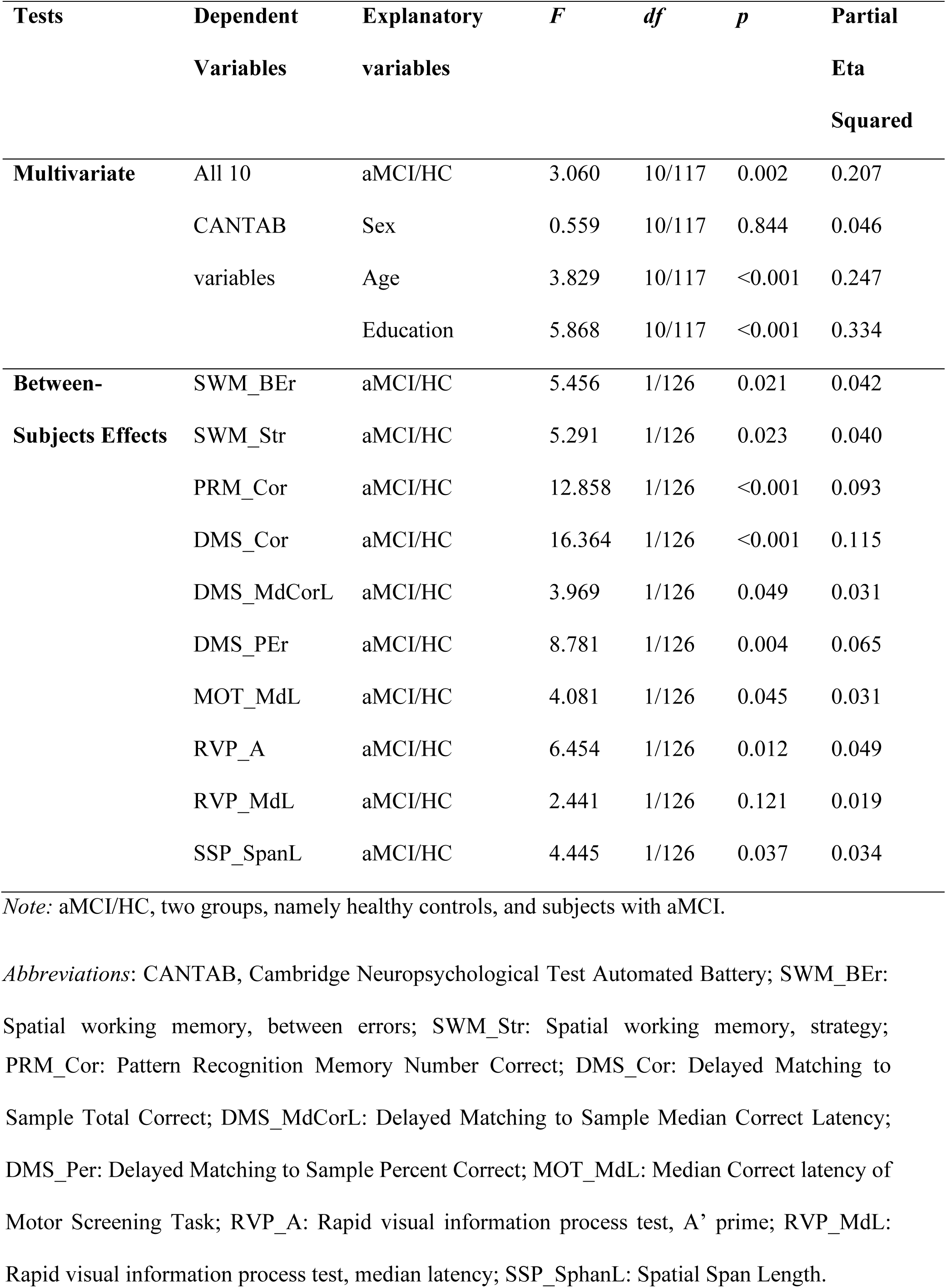
Results of multivariate GLM analysis which examines the association between the 10 Cambridge Neuropsychological Test Automated Battery (CANTAB) test scores and the clinical diagnosis of amnestic Mild Cognitive Impairment (aMCI)

| Tests | Dependent Variables | Explanatory variables | *F* | *df* | *p* | Partial Eta Squared |
| --- | --- | --- | --- | --- | --- | --- |
| **Multivariate** | All 10 | aMCI/HC | 3.060 | 10/117 | 0.002 | 0.207 |
|  | CANTAB | Sex | 0.559 | 10/117 | 0.844 | 0.046 |
|  | variables | Age | 3.829 | 10/117 | <0.001 | 0.247 |
|  |  | Education | 5.868 | 10/117 | <0.001 | 0.334 |
| **Between-Subjects Effects** | SWM_BEr | aMCI/HC | 5.456 | 1/126 | 0.021 | 0.042 |
|  | SWM_Str | aMCI/HC | 5.291 | 1/126 | 0.023 | 0.040 |
|  | PRM_Cor | aMCI/HC | 12.858 | 1/126 | <0.001 | 0.093 |
|  | DMS_Cor | aMCI/HC | 16.364 | 1/126 | <0.001 | 0.115 |
|  | DMS_MdCorL | aMCI/HC | 3.969 | 1/126 | 0.049 | 0.031 |
|  | DMS_PEr | aMCI/HC | 8.781 | 1/126 | 0.004 | 0.065 |
|  | MOT_MdL | aMCI/HC | 4.081 | 1/126 | 0.045 | 0.031 |
|  | RVP_A | aMCI/HC | 6.454 | 1/126 | 0.012 | 0.049 |
|  | RVP_MdL | aMCI/HC | 2.441 | 1/126 | 0.121 | 0.019 |
|  | SSP_SpanL | aMCI/HC | 4.445 | 1/126 | 0.037 | 0.034 |

*Note:* aMCI/HC, two groups, namely healthy controls, and subjects with aMCI.
*Abbreviations:* CANTAB, Cambridge Neuropsychological Test Automated Battery; SWM\_BEr: Spatial working memory, between errors; SWM\_Str: Spatial working memory, strategy;
PRM\_Cor: Pattern Recognition Memory Number Correct; DMS\_Cor: Delayed Matching to Sample Total Correct; DMS\_MdCorL: Delayed Matching to Sample Median Correct Latency; DMS\_Per: Delayed Matching to Sample Percent Correct; MOT\_MdL: Median Correct latency of Motor Screening Task; RVP\_A: Rapid visual information process test, A' prime; RVP\_MdL: Rapid visual information process test, median latency; SSP\_SphanL: Spatial Span Length.

**Table 3.** Model-generated estimated marginal mean (SE) values obtained by GLM analysis after adjusting for sex, age, and education

| **Variables** | **HC** | **aMCI** |
| --- | --- | --- |
| SWM_BEr | 49.02 (1.84) | 55.27 (1.82) |
| SWM_Str | 39.19 (0.42) | 39.59 (0.41) |
| PRM_Cor | 86.60 (1.30) | 79.80 (1.29) |
| DMS_Cor | 81.82 (1.23) | 75.17 (1.12) |
| DMS_MdCorL | 3735.5 (144.5) | 4154.9 (143.4) |
| DMS_PEr | 0.136 (0.019) | 0.216 (0.019) |
| MOT_MdL | 901.44 (31.41) | 993.85 (31.16) |
| RVP_A | 0.867 (0.007) | 0.841 (0.007) |
| RVP_MdL | 485.7 (16.4) | 523.0 (16.3) |
| SSP_SpanL | 5.20 (0.128) | 4.81 (0.126) |

*Abbreviations:* HC: Healthy controls; aMCI: Amnesic mild cognitive impairment; SWM\_BEr: Spatial working memory between errors; SWM\_Str: Spatial working memory strategy; PRM\_Cor: Pattern Recognition Memory Number Correct; DMS\_Cor: Delayed Matching to Sample Total Correct; DMS\_MdCorL: Delayed Matching to Sample Median Correct Latency; DMS\_Per: Delayed Matching to Sample Percent Correct; MOT\_MdL: Median Correct latency of Motor Screening Task; RVP\_A: Rapid visual information process test A' prime; RVP\_MdL: Rapid visual information process test, median latency; SSP\_SphanL: Spatial Span Length.

### Results of supervised and unsupervised machine learning

Using supervised machine learning (binary regression analysis and neural networks) we have delineated the best predictors and features of aMCI versus controls. **Table 4** depicts the findings of an automatic logistic regression (forward stepwise with p-to-enter of 0.05) with the control group as the reference and aMCI as the dependent variable, whilst allowing for the effects of the variables age, sex, and education. The most significant features of aMCI were PRM_Cor and DMS_Cor (Nagelkerke pseudo-R2 = 0.362; χ2 = 41.51, df = 2, p < 0.001). With a sensitivity of 68.2% and a specificity of 73.8%, 71.0% of all subjects could be correctly classified. Age, sex, and education were not significant discriminatory variables.

**Table 4.** Results of binary logistic regression with amnestic mild cognitive impairment (aMCI) as dependent variable and the two Cambridge Neuropsychological Test Automated Battery (CANTAB) subdomain scores as explanatory variables

| Variable | B | S.E. | Wald | df | p | OR | 95% CI for OR |
| --- | --- | --- | --- | --- | --- | --- | --- |
| PRM_Cor | -0.829 | 0.258 | 10.300 | 1 | 0.001 | 0.437 | 0.263-0.724 |
| DMS_Cor | -1.066 | 0.285 | 13.951 | 1 | <0.001 | 0.344 | 0.197-0.603 |

*Abbreviations:* OR: Odds ratio with 95 confidence intervals (CI); PRM\_Cor: Pattern Recognition
Memory Number Correct; DMS\_Cor: Delayed Matching to Sample Total Correct.

**Table 5** shows the results of MLP neural network analysis that used the CANTAB scores as input variables to predict either aMCI or the normal control group (output variables). An automated architecture training network comprised of two hidden layers was used: layer 1 had four units (hyperbolic tangent as the activation function) and layer 2 had three units (softmax as the activation function). The cross-entropy errors were significantly lower in the testing than in the training set. Inaccurate prediction rates in the training (30.5%), testing (25.0%), and holdout (29.2%) samples were rather consistent, showing that the model is not overtrained. **Table 6** shows that with a sensitivity of 77.3 percent, specificity of 65.4 percent, and an AUR ROC of 0.760, 70.8 percent of all subjects were properly identified. The importance of the input variables is depicted in **Figure 1** whereby the DSM_Cor and PRM_Cor were seen to be the most important determinants of the model’s predictive capacity. These two variables are then followed at a distance by DMS_Per and SWM_Ber, and again at a distance by the RVP scores and SWM_Str.

**Figure 1.**
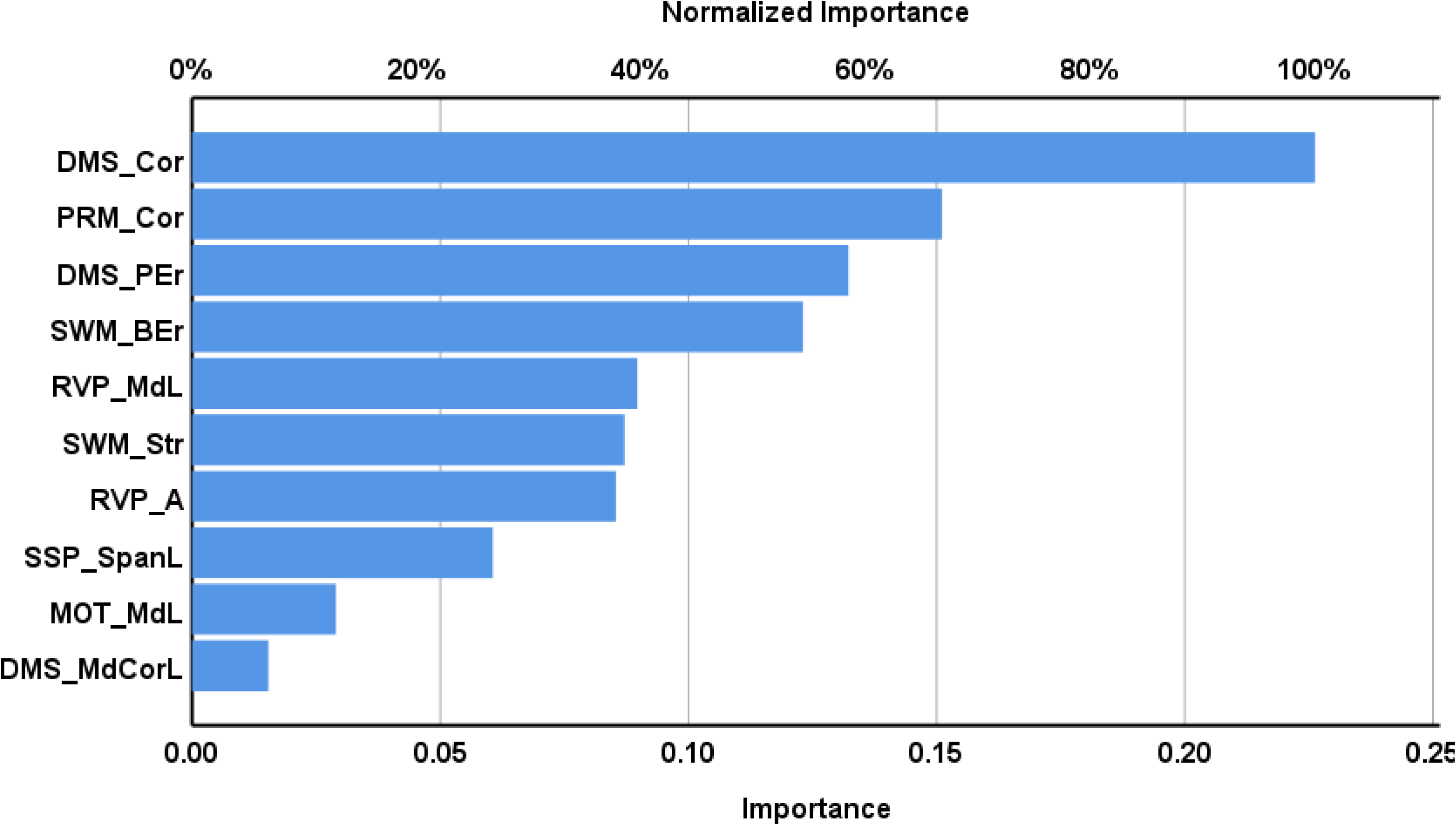
Neural network importance chart showing the relative and normalized importances of the CANTAB as input variables predicting amnestic mild cognitive impairment versus healthy controls (output variables). DMS_Cor: Delayed Matching to Sample Total Correct; PRM_Cor: Pattern Recognition Memory Number Correct; DMS_Per: Delayed Matching to Sample Percent Correct; SWM_BEr: Spatial working memory, between errors; RVP_MdL: Rapid visual information process test, median latency; SWM_Str: Spatial working memory, strategy; RVP_A: Rapid visual information process test, A’ prime; SSP_SphanL: Spatial Span Length; MOT_MdL: Median Correct latency of Motor Screening Task; DMS_MdCorL, Delayed Matching to Sample Median Correct Latency.

**Table 5.** Results of Multilayer Perceptron Neural Network analysis with amnestic mild cognitive impairment (aMCI) and healthy controls (HC) as output variables and the 10 Cambridge Neuropsychological Test Automated Battery (CANTAB) subdomain scores as explanatory variables.

| aMCI versus<br>HC | Samples | Cross<br>entropy<br>error term | Incorrect<br>predictions | Observed | Predicted<br>(sens/spec) | AUC<br>ROC |
| --- | --- | --- | --- | --- | --- | --- |
| 10 CANTAB<br>subdomains | Training | 12.117 | 30.5% | HC | 80.6% | 0.760 |
|  |  |  |  | aMCI | 57.1% |  |
|  | Testing | 4.596 | 25.0% | HC | 83.3% |  |
|  |  |  |  | aMCI | 66.7% |  |
|  | Holdout | - | 29.2% | HC | 77.3% |  |
|  |  |  |  | aMCI | 65.4% |  |

**Table 6.** Results of multiple regression analysis with Thai Mini-Mental State Examination (TMSE) and Thai Montreal Cognitive Assessment (MoCA-Thai) as the outcome variables and the Cambridge Neuropsychological Test Automated Battery (CANTAB) subdomain scores as explanatory variables

| Dependent variables | Explanatory variables | $\beta$ | t | p | F model | df | p | R<sup>2</sup> |
| --- | --- | --- | --- | --- | --- | --- | --- | --- |
| #1. TSME | PRM_Cor | 0.338 | 4.26 | <0.001 | 26.81 | 3/130 | <0.001 | 0.388 |
|  | RVP_A | 0.255 | 3.24 | 0.002 |  |  |  |  |
|  | Education | 0.203 | 2.44 | 0.016 |  |  |  |  |
| #2. MoCA-Thai | DMS_Cor | 0.403 | 4.89 | <0.001 | 24.370 | 3/130 | <0.001 | 0.365 |
|  | PRM_Cor | 0.182 | 2.23 | 0.027 |  |  |  |  |
|  | RVP_MdL | -0.169 | -2.15 | 0.033 |  |  |  |  |

*Abbreviations:* PRM\_Cor: Pattern Recognition Memory number correct; RVP\_A': Rapid Visual Information process test, A' prime; DMS\_Cor: Delayed Matching to Sample total correct; RVP\_MdL: Rapid Visual Information Process latency median.

K-means cluster analysis using the 10 CANTAB scores discovered 2 clusters with a good silhouette measure of cohesion and separation of 0.51. The first cluster comprised 94 subjects, namely 36 aMCI patients and 58 controls, and a second cluster showing impairments in all 10 neurocognitive tests (except DMS_MdCorL) comprised 37 subjects, namely 30 aMCI patients and 7 controls (χ2 = 19.44, df = 1, p < 0.001). The clustered bar graph displaying the mean (SE) CANTAB test scores in both clusters can be seen in **Figure 2**. These were no significant differences in sex distribution between both groups, but mean (SD) age (75.5 ±5.9 versus 69.6 ±6.2 years) was higher (F=24.06, df=1/129, p<0.001) and mean (SD) years of education (8.1 ±4.6 versus 13.2 ±13.2) lower (F=30.9, df=1/129, p<0.001) in the second cluster. Nevertheless, multivariate GLM analysis revealed that the effect size of the clusters (0.540) was much greater than that of age (0.140), education (0.284), and sex (0.051).

**Figure 2.**
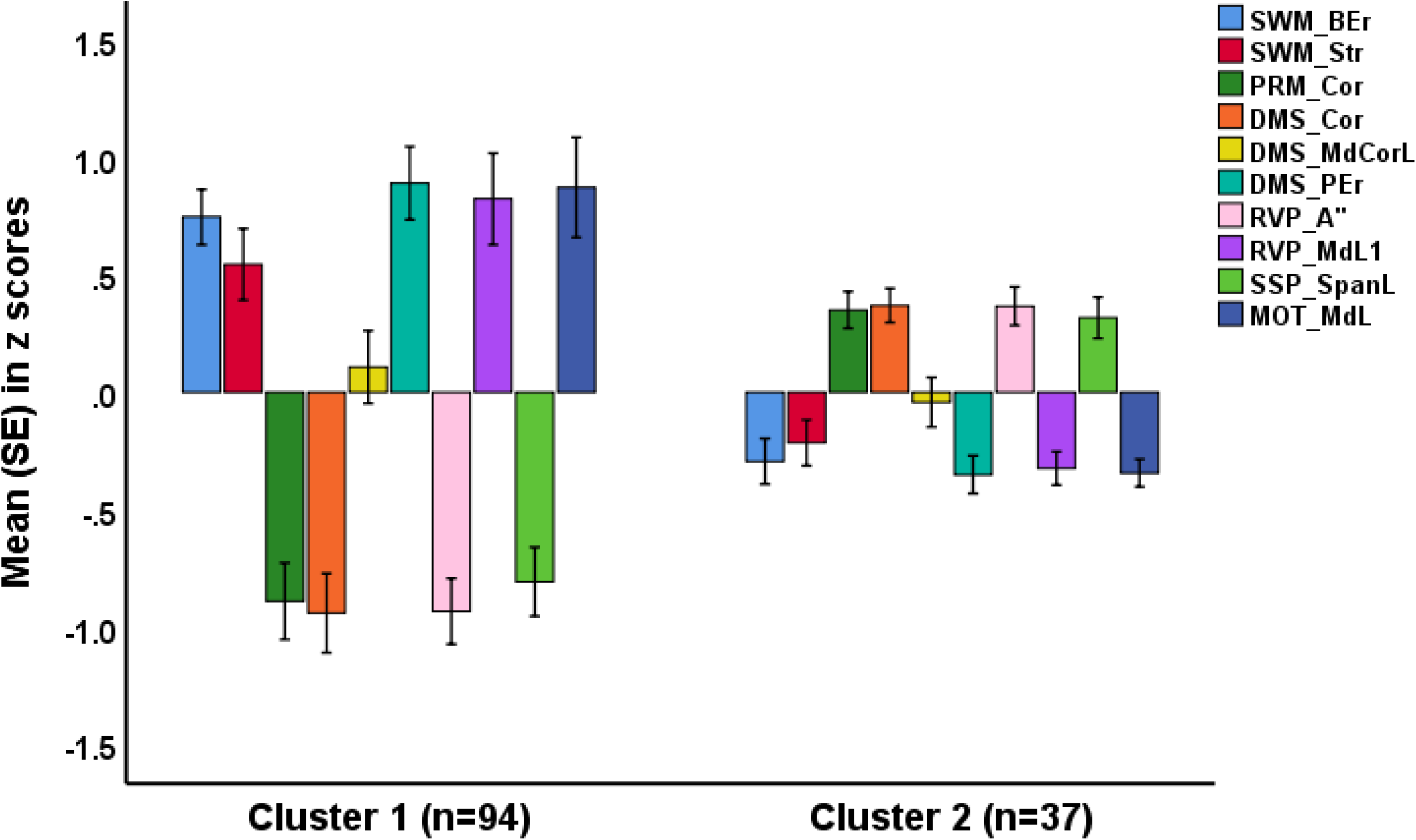
Clustered bar graph showing the z scores of the subdomains of 10 Cambridge Neuropsychological Test Automated Battery (CANTAB) test scores in two cluster-analysis-generated classes.

### Results of regression and PLS-path analysis

To delineate the CANTAB features that best capture the MoCA and TMSE scores in aMCI, we performed multiple regression analysis with MoCA/TMSE scores as dependent variables and PLS analysis with a latent vector extracted from MoCA, TMSE, and the diagnosis aMCI as output variable. **Table 6**, regression #1 shows that PRM_Cor, RVP_A and education explained 38.8% of the variance in TSME (all positively), while regression #2 reveals that 36.5% of the MoCA score’s variance was explained by DMS_Cor and PRM_Cor (both positively) and RVP_MdL.

The final PLS model is shown in **Figure 3**. The outcome indicator is a latent vector extracted from MoCA, TMSE and the aMCI diagnosis versus controls (dubbed as CLINICS). All CANTAB test scores, age, sex, and education were input indicators. Non-significant indicators were deleted from the final model. With SRMR = 0.031, the model in **Figure 3** exhibits an adequate model fit. The construct reliability of the latent vector was more than efficient with an AVE value > 0.759, composite reliability of 0.94 and significant loadings > 0.792 (at p<0.0001). PLS Predict revealed that this latent vector was not misspecified as a reflective model and blindfolding revealed an appropriate construct cross-validated redundancy. The indicators’ Q2 predict scores were all positive, indicating that they outperformed the naivest benchmark. We found that SWM_Ber, RVP_MdL and DMS_Cor explained 29.0% of the variance in the outcome variable. The model shows that age and education explained 11.1% of the variance in SWM_Ber, and that education accounts for 3.8% and 8.3% of the variances in DMS_Cor and RVP_MdL, respectively. Furthermore, the specific, indirect effects revealed that all pathways between input indicators and the latent vector were significant, and that SWM_Ber, RVP_MdL, and DMS_Cor mediated the effects of age and education on CLINICS. On the other hand, MOT_MdL had no significant impact on the outcome indicator while age and education explained 9% of the variance in MOT_MdL.

**Figure 3.**
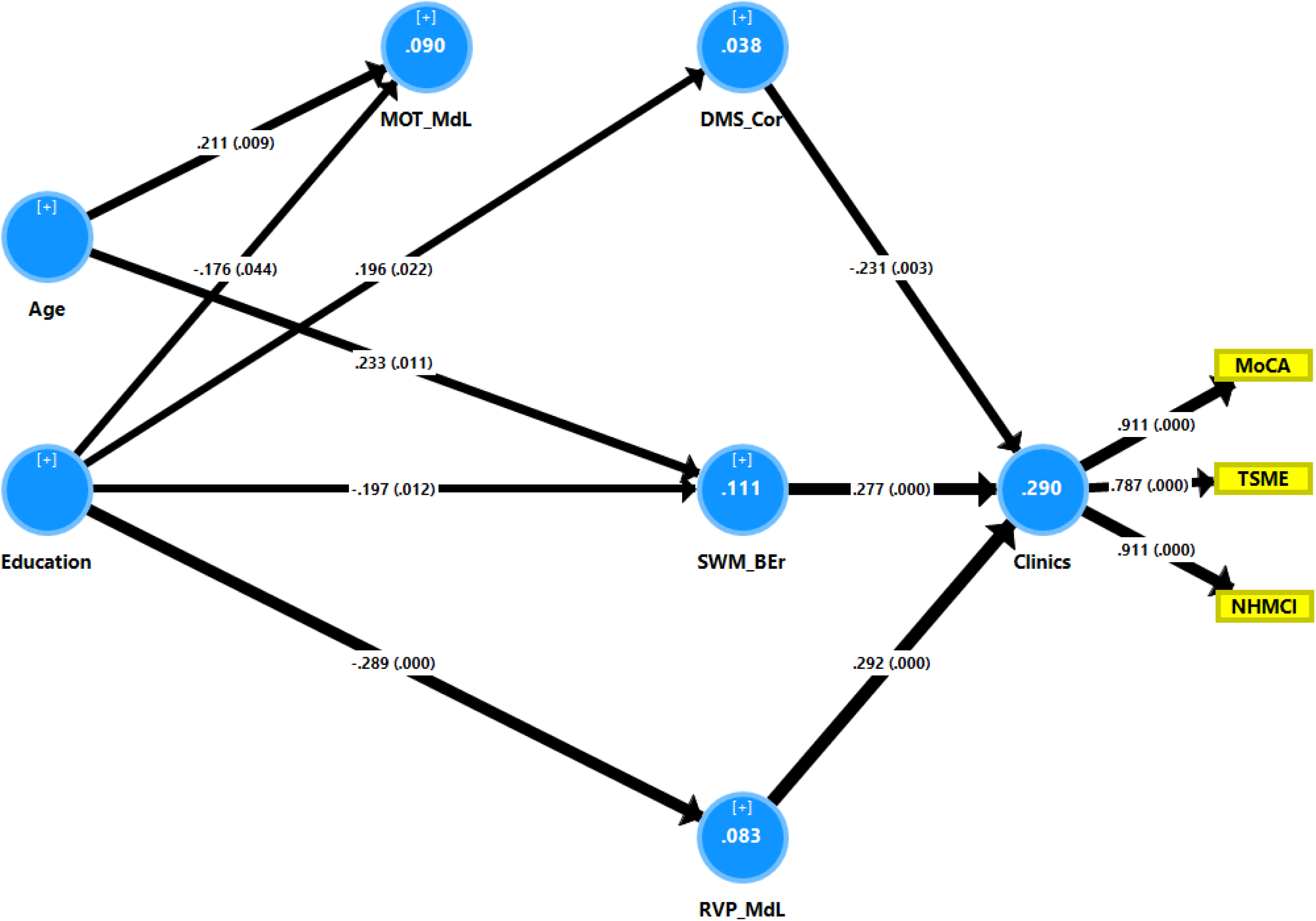
Results of complete partial least squares analysis performed on 5000 bootstrap samples. Path coefficients and loadings (with P values) are shown. White figures within the circles indicate percentage of variance explained. Sex, age and education are input variables and clinical data are output variables with cognitive data mediating the effects of the input on the output variables. CLINICS: a latent vector extracted from TSME (Thai Mini-Mental State Examination), MoCA (Montreal Cognitive Assessment-Basic) and diagnosis of amnestic Mild Cognitive Impairment (HCMCI); DMS_Cor: Delayed Matching to Sample, Total Correct; MOT_MdL: Median Correct latency of Motor Screening Test; SWM_Ber: Spatial Working Memory, Between errors; RVP_MdL: Rapid Visual information Process test, Median Latency.

## Discussion

The first major finding of this study is that, in accordance with our a priori hypothesis, the clinical diagnosis of aMCI is validated by CANTAB tests including spatial working, pattern recognition, short-term visual recognition and visuospatial working memory as well as rapid visual information processing. These findings are in line with previous reports that impairments in executive cognition (such as Spatial Working Memory Strategy and Spatial Working Memory Between Errors) are relatively common among older adults with mild cognitive impairment (Brandt et al., 2009; Zheng et al., 2012). Our results agree with an article published by Alichniewicz et al. (2010) showing that aMCI patients had reduced skills in visual memory, sustained attention, and spatial planning memory, compared to healthy subjects. Previous research has discovered that aMCI is characterized by deficiencies in Spatial Working and pattern Recognition Memory and Delayed Matching to Sample (Jirayucharoensak et al., 2019). On the other hand, MCI patients performed similarly to control subjects on visual recognition memory tests (Delayed Matching to Sample, Pattern Recognition Memory, and Spatial Working Memory) (Barbeau et al., 2004). Interestingly, the present study indicated that aMCI is not only accompanied by memory dysfunctions but also by a functional deficit in motor abilities (as assessed with the Motor Screening Task), which is a feature of dementia (Schonfeld et al., 2021).

The second major finding is that using machine learning approaches we were able to delineate the key aMCI features, namely impairments in pattern recognition and short-term visual recognition memory, whilst deficits in spatial working memory and rapid visual processing are other, albeit somewhat less important features of aMCI. Although education and age are strongly associated with the CATBAB test results, the associations between the clinical aMCI diagnosis and CANTAB tests are more important than with education and age (results of binary regression analysis). This data indicates that memory impairments coupled with executive dysfunctions are the primary features of aMCI. Such data may suggest that aMCI is accompanied by more specific dysfunctions in brain circuits or brain structures. For example, Reinvang et al. (2012) found that the volume of the dorsolateral prefrontal cortex, fusiform gyrus, and posterior parietal cortex strongly correlated with working memory in MCI (Ilardi et al., 2021).

Nevertheless, the effect sizes of the associations between the CANTAB tests and aMCI are modest. Thus, GLM analysis indicated that the shared variance between the CANTAB tests and the clinical diagnosis aMCI was always <11.5% (DMS_Cor) and sometimes as low as 3.1% (DMS_MdCor). Moreover, both logistic regression and neuronal networks showed that, using the CANTAB test scores as input variables, only 70-76% of all subjects were correctly classified as either aMCI subjects or controls. Furthermore, the CANTAB test results explain only 29.1%- 38.8% of the variance in the total MoCA and TMSE scores, which may be used as external validating criteria for the aMCI diagnosis.

The third major finding of this study is that the K-means cluster analysis performed on the CANTAB test results delineated a well-validated cluster of subjects with more severe neurocognitive impairments and that this cluster comprised 45.5% of the subjects with aMCI while 54.5% of aMCI subjects were allocated to the control group. In addition, up to 10.8% of our controls were - based on the CANTAB test results - allocated to the group with neurocognitive impairments. As such, it appears that the clinical aMCI group, as diagnosed using Petersen’s criteria, is a heterogeneous group, which is additionally overinclusive because many older adults are wrongly classified as aMCI. It follows that new diagnostic criteria of aMCI should be based on more restrictive criteria comprising impairments in different CANTAB subdomain scores. In this regard, it is important to note that, using another supervised learning technique (such as the soft independent modeling of class analogy, or SIMCA), we showed that aMCI is a heterogeneous group based on Consortium to Establish a Registry for Alzheimer’s Disease (CERAD) probes of episodic and semantic memory and that many people who were diagnosed with aMCI actually belong to the control group, while a few were allocated to the AD sample (Tangwongchai et al., 2018).

Another study was unable to validate the aMCI study group using nearest neighbour analyses (Maes & Tangwongchai, 2021). The latter study computed a composite score using CERAD probe scores, behavioural changes, deficits in ADL, the ApoE4 allele, white blood cells, serum albumin, folate, the atherogenic index of plasma and fasting blood glucose (FBG), as well as comorbidities such as type 2 diabetes mellitus and hypertension as modelling variables (Maes & Tangwongchai, 2021). Based on those combined neurocognitive, behavioural, and biomarker features, these authors were able to construct a subgroup of people who were placed at an intermediate stage between controls and AD patients. Again, this subgroup was more restrictive than the clinical aMCI group as defined with the Petersen criteria and in addition displayed memory, executive, language, Behavioral, ADL and biomarker alterations clearly separating this group from controls and AD patients (Maes & Tangwongchai, 2021).

Collectively, the results of the current study and those of Maes and Tangwongchai (2021) and Tangwongchai et al. (2018) show that the clinical diagnosis of aMCI is overinclusive because too many healthy controls are allocated to this diagnostic group. Furthermore, it appears that focusing too much on the memory disorders and excluding mild impairments in ADL and behaviours from the aMCI diagnostic criteria further complicates the classification of individuals belonging to the intermediate group. In summary, Petersen’s aMCI diagnosis could not be validated and may be better represented by a more restrictive cluster of subjects defined by mild impairments in memory and executive functions, as well as behavioral and biomarker features. The latter findings are further underscored by recent findings that aMCI is characterized by increased oxidative stress and lowered antioxidant defences (Nantachai et al., 2022).

There are some limitations that should be considered regarding the results of this study. First, this study was limited to patients with aMCI and did not distinguish between additional subgroups such as single-domain and multi-domain aMCI, which may possess different underlying etiologies and give different outcomes (Brambati et al., 2009; Petersen, 2007). Second, future studies should incorporate CANTAB and CERAD test findings with measures of memory, executive functions, ADL, behavior, and biomarkers including nitro-oxidative stress biomarkers.

## Conclusions

Deficits in memory including spatial working memory, pattern recognition memory, short-term visual recognition memory, and visuospatial working memory capacity as well as rapid visual information processing are key features of aMCI as diagnosed with Petersen’s criteria. Nevertheless, the results of machine learning techniques show that the associations between aMCI and CANTAB tests have a small effect size, and that the clinical diagnosis of aMCI is overinclusive because too many healthy controls are allocated to this diagnostic group.

## Disclosure

The authors report no conflict of interest with any commercial or other association in connection with the submitted article.

## Authorship Contributions

All authors contributed to the writing up of the article.

## Data Availability Statement

The dataset generated during and/or analysed during the current study will be available from the corresponding author MM upon reasonable request and once the dataset has been fully exploited by the authors.

Research Involving Human Participants

Approval for the study was obtained from the Institutional Review Board of Chulalongkorn University, Bangkok, Thailand (No. 359/56).

## Informed Consent

All controls and aMCI subjects gave written informed consent before participation in our study.

## Data Availability

The dataset generated during and/or analyzed during the current study will be available from the corresponding author upon reasonable request and once the dataset has been fully exploited by the authors.

